# Effectiveness and Implementation Costs of School Share Tables in the Midwestern United States

**DOI:** 10.1101/2025.06.23.25330059

**Authors:** Melissa Pflugh Prescott, Trinity Allison, Stephanie Pike Moore, Gwendoline Balto, Matthew J Stasiewicz

## Abstract

**Objective:** To evaluate the impact of a train-the-trainer approach to share table implementation on school food recovery and to determine the maintenance costs of share table operation.

**Methods:** In 2022-2023, researchers collaborated with the Supplemental Nutrition Assistance Program-Education (SNAP-Ed) Programs in 6 states (Illinois, Indiana, Michigan, Minnesota, Ohio, Wisconsin) to implement and evaluate the integration of share tables into school meal programs. Each participating school (n= 41) submitted monthly cost surveys, which assessed the costs of share table eligible food and beverage items ordered and any costs associated with share table implementation. SNAP-Ed staff conducted bimonthly Share Table observations to assess student use of share tables, use of recovered food items (taken by students, re-serviced, donated, or discarded), and weight of recovered food items.

**Results:** An estimated 978 kg and $2,565 was recovered per school in the first year of share table operation. Each school expended an average of $4.15 over the school year to maintain share table operations, and only 1 school reported an increase in staff time.

**Conclusions:** These findings suggest that share tables are inexpensive to maintain while also improving access to healthy food and reducing food waste and school nutrition program food costs.

## Introduction

Despite 12.8% of Americans experiencing food insecurity^1^, food comprises 24% of municipal solid waste in landfills, making it the single largest landfilled material^2^.Food production and waste management contribute to a variety of environmental impacts, such as climate change, air pollutants, water scarcity, biodiversity loss, and soil and water quality degradation. For example, the World Wildlife Fund estimates that solid school food waste alone costs the earth 1.9 million metric tons of greenhouse gases and 20.9 billion gallons of water waste each year^3^. Climate change is a public health concern because certain groups, like children, the elderly, and communities of color, are less climate resilient and subsequently are more vulnerable to the negative health effects of climate change, resulting in disparate health outcomes^4–6^. In the interest of mitigating climate change, the U.S. Environmental Protection Agency (EPA) provides guidance on the sustainable use of food, proposing that food waste first be prevented and when this fails, food should be upcycled or donated^7^. A 2019 study identified school share tables as the most feasible school food recovery method for student plate waste.^8^ Share tables are places in school cafeterias where students can donate their unwanted school food items to be either eaten as second helpings by hungry students, donated to non-profits, or re-serviced (i.e. re-used in future meal services). Given that all foods sold in school meals are subject to rigorous nutrition standards, food recovered on share tables can be used to address food and nutrition insecurity, as well as climate change mitigation. In addition, the USDA allows re-serviced share table items to be a part of future reimbursable meals. For example, purchased yet uneaten apples from lunch on Monday can be resold at breakfast on Tuesday, and this same apple would be part of two reimbursable meals, allowing schools to concurrently reduce food costs and their environmental footprint.

While share tables provide a sound method for food recovery, they add additional steps to the lifespan of each food item-thereby introducing additional food safety risk.^9^ Even though food safety risks of share table operation should theoretically be the same for schools across the U.S., state-level policies vary on what is permitted to be collected and the methods of re-using unclaimed share table items (e.g. reservice, external donation). In 2021, qualitative study examined the share table food borne illness concerns of health inspectors and concluded that the risk of Norovirus via unwrapped apples was perceived as the most hazardous safety risk.^10^ Yet, simulation studies suggest that this risk is mitigated by student hand washing or sanitizing, and school food service staff can further mitigate norovirus risk by implementing Share-Only share tables, where students may place items on the share table but not take them off, and by washing recovered apples prior to re-serving in future meals.^11^ Another study concluded that share tables do not meaningfully reduce microbial quality of milk, but that school food service staff should return shared milk to refrigeration as soon as possible.^12^ Given the key role of food safety in share table operation, the USDA recommends that schools obtain approval from their local health department prior to share table operation.^13^

Given that the USDA allows share table operation in all school nutrition programs, widespread share table adoption has the capacity to mitigate climate change, cut school food costs, and make healthy foods available to vulnerable populations. In addition, estimating the costs of share table maintenance and potential to re-coup food costs through share table food recovery may encourage additional schools to adopt share tables. However, feasibility of share table operation may be improved by providing technical assistance for food safety and other aspects of implementation. The purpose of this study is to estimate the rate of school food and food cost recovery and the maintenance costs of share tables implemented using a train-the-trainer approach.

## Methods

In this non-randomized pilot study, researchers collaborated with the Supplemental Nutrition Assistance Program-Education (SNAP-Ed) Programs in 6 midwestern states (Illinois, Indiana, Michigan, Minnesota, Ohio, Wisconsin) to evaluate school share table implementation into school meal programs. SNAP-Ed programs work with schools with a high proportion of students who receive free or reduced priced meals to provide technical assistance to support the adoption and implementation of evidence-based strategies to improve access to healthy food and promote positive nutrition-related behaviors^13^. Methods used in this study were approved by the University of Illinois at Urbana-Champaign Institutional Review Board.

Recruitment took place in March 2022. Researchers worked with state-level SNAP-Ed administrators to recruit SNAP-Ed technical assistance providers who would then be asked to recruit schools with 50% free or reduced priced lunch eligibility in their region to participate in the pilot. The allocated grant funding provided resources to include up to 60 schools in the pilot. Census data was used to determine the maximum number of schools each state could recruit to reflect actual population distributions across the 6 state region. (IL: 14-15 schools, IN 7-8 schools, MI: 11-12 staff, MN: 6-7 schools, OH: 13-14 schools, and WI: 6-7 schools. A total of 52 schools were contacted, and 41 participated in the pilot. All participating SNAP-Ed staff were trained on share table food safety and operating procedures through a series of live webinars.

### Intervention

The intervention consisted of nutrition services staff trainings and share table operation technical assistance provided by local SNAP-Educators, as well as support in obtaining health department approval of share tables. Health departments differed as to what types of food items they permitted to be included on share tables and how unclaimed/leftover share table foods could be used (donated, re-serviced, or landfilled), with milk being the most likely item to be restricted from share tables or from being re-used via donation or re-serviced. Participating schools also had access to a Share Table Tool Kit, developed by [redacted], which consisted of standard operating procedures, signage, communication tools, food safety plans, and other resources to assist with share table operation. As an incentive for participating in the pilot, each participating school had the option to receive a Share Table Equipment Kit (consisting of a wheeled chrome-plated steel wire cart, washable baskets, 25-quart cooler, signs, and 300 count reusable ice cubes) and valued at $800 including shipping costs at the time of the intervention) or an $800 check to purchase their own share table supplies.

### Evaluation

Each participating school submitted monthly cost surveys, which assessed the quantities and price per unit of share table eligible food and beverage items ordered, as well as potential costs associated with Share Table implementation, such as staff-related expenses (overtime/increased work hours) and additional incurred operational costs such as additional ice packs and printed signs.

Bimonthly Share Table observations were conducted by SNAP-Ed staff, starting one month post share table implementation. SNAP-Ed staff were asked to observe all lunch periods using a structured observation protocol to document the number of eligible items placed on and removed from the share table, the quantity, weight and intended use of leftover items (re-service, donation, or discard). Some schools preferred their share table to focus on breakfast service, and observations of breakfast periods were made at those schools instead of at lunch.

Using information captured from both the structured observations as well as the cost surveys, the estimated weight (in kg) and cost (in U.S. dollars (USD)) was calculated for each unit of food recovered (e.g., taken during meal period, re-serviced, donated) and those that were discarded. To make valid comparisons across states with varying degrees of school participation, the average weight and cost of foods recovered were calculated across all meal periods observed. In addition to examining the magnitude of foods recovered, we also examined the average proportion of items that were recovered and discarded across each observation.

### Analysis

Bivariate analyses using ANOVA were used to examine descriptive differences between states with respect to average number of observations, average number of meal periods observed, average number of students participating in school lunches, overall school enrollment as well as the average weight and cost of foods recovered and discarded for each meal period observed. Fisher’s exact tests were used to examine differences in participating school characteristics across states including school type, share table type, or reporting additional costs incurred.

To estimate the potential impact of this regional intervention across the school year, we estimated the total potential weight and costs of foods recovered and discarded by taking the averages for each meal period observed and multiplying them by the regular number of meal periods per day and then by the minimum number of school days legally required for each state for each school. We fit a series of Poisson regression models using the PROC GENMOD procedure in SAS (v9.4) using each weight and cost recovery category as the dependent variables and state as the independent, explanatory variable. We calculated the least squares means for each of the outcomes to estimate the mean food weight and cost recovery categories using the inverse link function to transform estimates back to their original scale. We computed 95% prediction intervals (PI) for each variable and exponentiating them to transform the intervals to the original scale. These provided state-specific mean food weights and costs anticipated for individual schools in each state. To calculate estimates at the regional level, state-level estimates were weighted by the proportion of participating schools in each state and then summing the weighted mean estimates across all states.

## Results

Share-and-take share tables, where students can freely donate food and take donated food off the share table during the meal period, were the dominate share table type (80.5%) across the region (Table 1). Elementary schools (67.5%) were the most prevalent participating school type, and 85.4% of schools opted to receive the share table kit instead of the check incentive. Nine schools (22.0%) reported additional material costs of operating a share table beyond the pilot participation incentive. Of these, only four schools reported actual dollar amounts for additional costs and indicated they spent a total of $17 to $69, or $4.15 on average across all 41 participating schools, on additional materials such as shelf liners, signage, and ice sheets over the course of their participation. Notably, 75.0% of schools with additional costs opted to receive an $800 incentive rather than the share table materials. Only one school reported needing additional staff overtime of 1.5-2 hours per day for cart maintenance. This suggests that there are typically minimal day-to-day costs for share table operation. There were no significant differences in type of share table, school level, incentive preference, or additional costs across states.

**Table 1.**
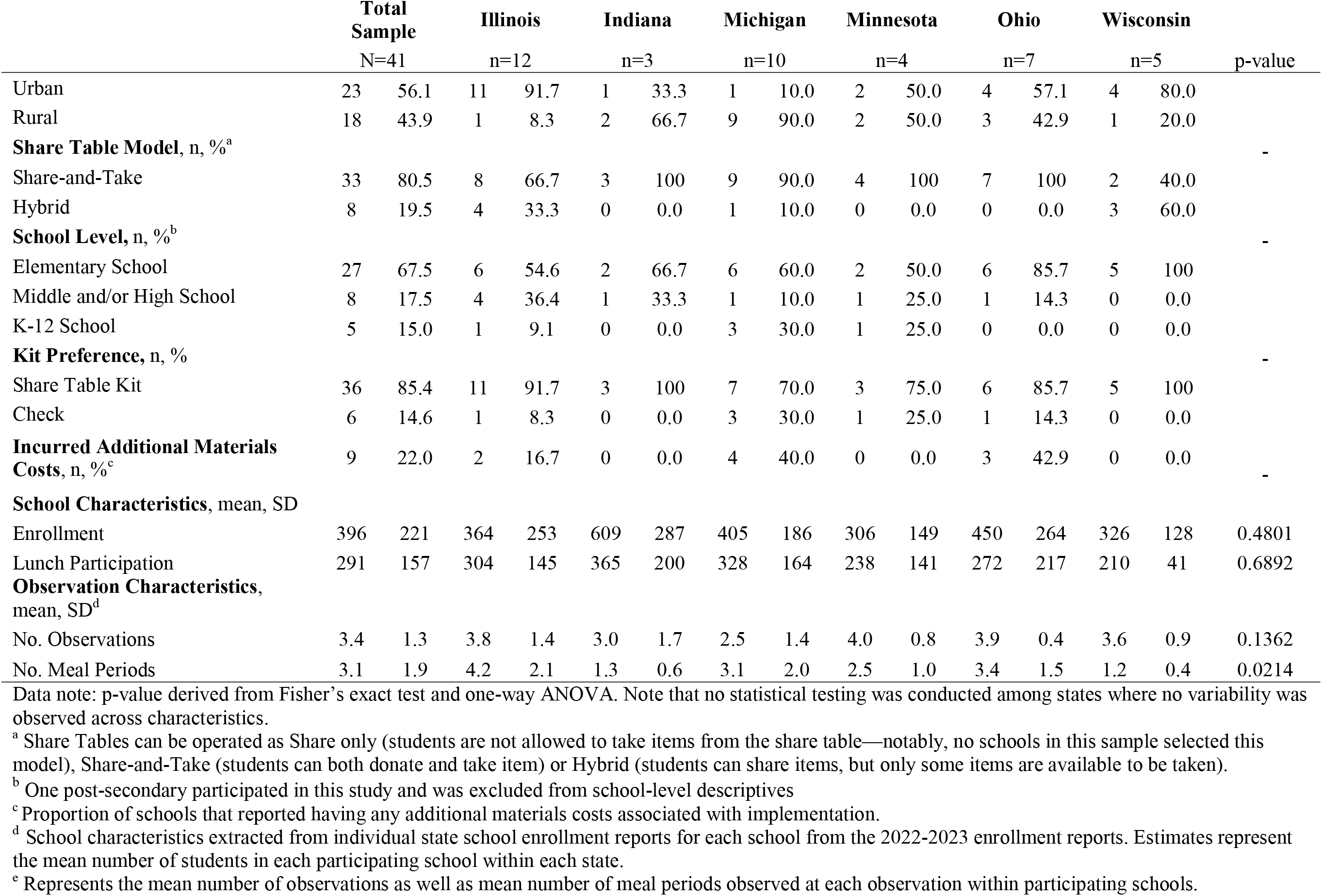
Regional and State-Level Characteristics of Share Table Implementation, 2022-2023 School Year.

Overall, there was a total of 140 observations across 41 participating schools (mean= 3.4 per school SD: 1.3)), a total of 426 lunch (or breakfast) periods observed (mean=3.1 per school per observation, SD: 1.9) (Table 2). Participating schools had approximately 16,242 enrolled students (mean=396.1, SD: 221.0). The combined average daily participation rate across the participating schools suggests that approximately 11,938 students had access to the piloted share tables each school day (mean=291.2 per school, SD: 156.5). There were statewide differences in the number of meal periods observed for each observation with Illinois having the greatest average number of meal periods observed (4.2; SD: 2.1) and Wisconsin (1.2; SD: 0.4)) and Indiana (1.3; SD: 0.6) having the fewest number of meal periods observed, on average.

**Table 2.** Mean Share Table Food Recovery Metrics Across School Observations (n=140), Per School.

|  | Observed Food Waste Recovery by Weight (kg) |  |  |  |  |  |  |  |  |  |  |  | Observed Discard Weight (kg) |  |  |
| --- | --- | --- | --- | --- | --- | --- | --- | --- | --- | --- | --- | --- | --- | --- | --- |
|  | Total |  |  | Taken |  |  | Reserviced |  |  | Donated |  |  |  |  |  |
|  | Total | Mean | SD | Total | Mean | SD | Total | Mean | SD | Total | Mean | SD | Total | Mean | SD |
| **Regional Total** | 708.3 | 2.1 | 2.3 | 465.7 | 1.5 | 1.8 | 149.8 | 0.4 | 0.7 | 92.8 | 0.2 | 0.6 | 43.2 | 0.1 | 0.2 |
| **State Total** |  |  |  |  |  |  | ** |  |  |  |  |  |  |  |  |
| Illinois | 329.4 | 1.6 | 1.1 | 223.7 | 1.0 | 0.9 | 70.3 | 0.4 | 0.5 | 35.4 | 0.2 | 0.4 | 8.4 | 0.1 | 0.1 |
| Indiana | 63.8 | 5.4 | 4.7 | 42.0 | 3.5 | 3.4 | 21.8 | 1.9 | 1.4 | 0.0 | - | - | 1.0 | 0.1 | 0.1 |
| Michigan | 136.4 | 2.6 | 3.1 | 54.8 | 1.8 | 2.8 | 26.6 | 0.3 | 0.5 | 55.1 | 0.6 | 1.0 | 12.0 | 0.1 | 0.2 |
| Minnesota | 30.9 | 1.1 | 0.9 | 24.9 | 0.9 | 1.0 | 6.0 | 0.1 | 0.1 | 0.0 | - | - | 0.2 | 0.0 | 0.0 |
| Ohio | 103.7 | 1.3 | 1.1 | 84.2 | 1.1 | 0.9 | 17.1 | 0.2 | 0.2 | 2.4 | 0.0 | 0.1 | 17.3 | 0.2 | 0.1 |
| Wisconsin | 44.1 | 2.2 | 1.5 | 36.2 | 1.8 | 1.3 | 7.9 | 0.4 | 0.2 | 0.0 | - | - | 4.3 | 0.2 | 0.3 |
| | Observed Food Waste Recovery by Cost Savings (USD/\$) | | | | | | | | | | | | Observed Discard Cost (USD/\$) | | |
|  | Total |  |  | Taken |  |  | Reserviced |  |  | Donated |  |  |  |  |  |
|  | Total | Mean | SD | Total | Mean | SD | Total | Mean | SD | Total | Mean | SD | Total | Mean | SD |
| **Regional Total** | 1867 | 5.36 | 4.98 | 1163 | 3.61 | 4.07 | 484 | 1.14 | 1.71 | 221 | 0.62 | 1.35 | 114 | 0.27 | 0.43 |
| **State Total** |  |  |  |  |  |  |  |  |  |  |  |  |  |  |  |
| Illinois | 937 | 4.81 | 2.88 | 580 | 2.76 | 2.31 | 251 | 1.39 | 1.85 | 106 | 0.66 | 1.29 | 21 | 0.14 | 0.35 |
| Indiana | 104 | 8.70 | 7.35 | 63 | 5.21 | 4.55 | 41 | 3.49 | 3.07 | 0 | - | - | 3 | 0.23 | 0.39 |
| Michigan | 318 | 6.98 | 7.83 | 138 | 4.73 | 6.98 | 80 | 0.69 | 1.70 | 101 | 1.57 | 2.03 | 30 | 0.24 | 0.29 |
| Minnesota | 65 | 2.25 | 1.94 | 52 | 1.99 | 2.10 | 13 | 0.26 | 0.28 | 0 | - | - | 1 | 0.02 | 0.02 |
| Ohio | 330 | 4.14 | 3.56 | 24 | 3.12 | 2.44 | 75 | 0.78 | 1.04 | 14 | 0.24 | 0.64 | 51 | 0.64 | 0.65 |
| Wisconsin | 113 | 5.67 | 3.21 | 89 | 4.44 | 2.77 | 25 | 1.23 | 1.04 | 0 | - | - | 8 | 0.41 | 0.52 |

Data note: Mean weight (kg) and cost estimate (USD, or U.S. Dollars) were calculated across each meal period during each observation across participating schools and represents the mean per school per meal period; p-value derived from one-way ANOVA test.

A total of 708.3 kg of food was recovered across 140 observations representing $1,867 in item costs. Across each school, the average weight in food recovered per meal period was 2.1 kg (SD: 2.3) and the average cost of foods recovered for each meal period was $5. There were no differences across states with respect to weight and costs of food recovered or discarded with the exception of the weight of reserviced food. In Indiana, there was an average of 1.9 kg (SD: 1.4) of food reserviced observed across each meal period which was greater than states like Minnesota where only 0.1 kg (SD: 0.1) of food items were reserviced.

Figure 1 illustrates differences in the proportion of weight of food recovered and discarded across each state. Minnesota had the highest average proportion of taken foods, with 82.1%, (95% CI: 66.2, 98.0) of the total weight of their recovered food being taken by students during meal periods. This is significantly different from Illinois (45.4%, 95% CI: 34.9, 55.8), who had the lowest share of food taken by weight. Indiana had the highest average proportion of re-serviced foods (41.0%, 95% CI: 20.9, 61.1)), while Ohio had significantly fewer re-serviced foods (7.5%, 95% CI: 1.1, 13.8). Only three states reported donating any foods recovered from share tables, with Michigan having the greatest proportion of donations by weight (16.7%, 95% CI: 5.7, 26.5). Ohio had the greatest proportion, on average, of food discarded by weight (23.8%, 95% CI: 12.8, 34.9), which was significantly higher than most other participating states. As shown in Figure 2, these same patterns held when examining the costs of food recovered across states.

**Figure 1.**
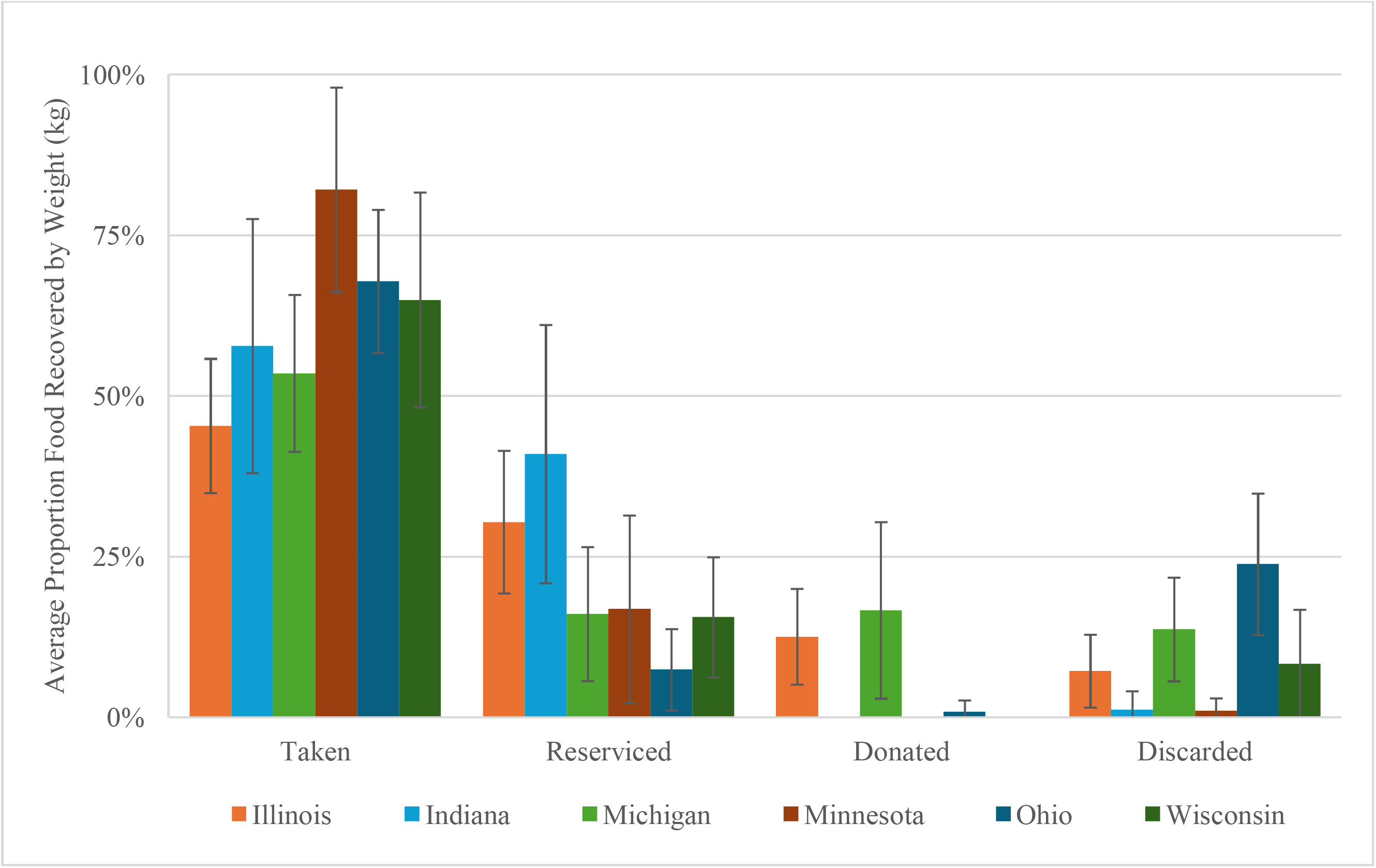
Weight of foods placed on share tables during observation meals by final use and state

**Figure 2.**
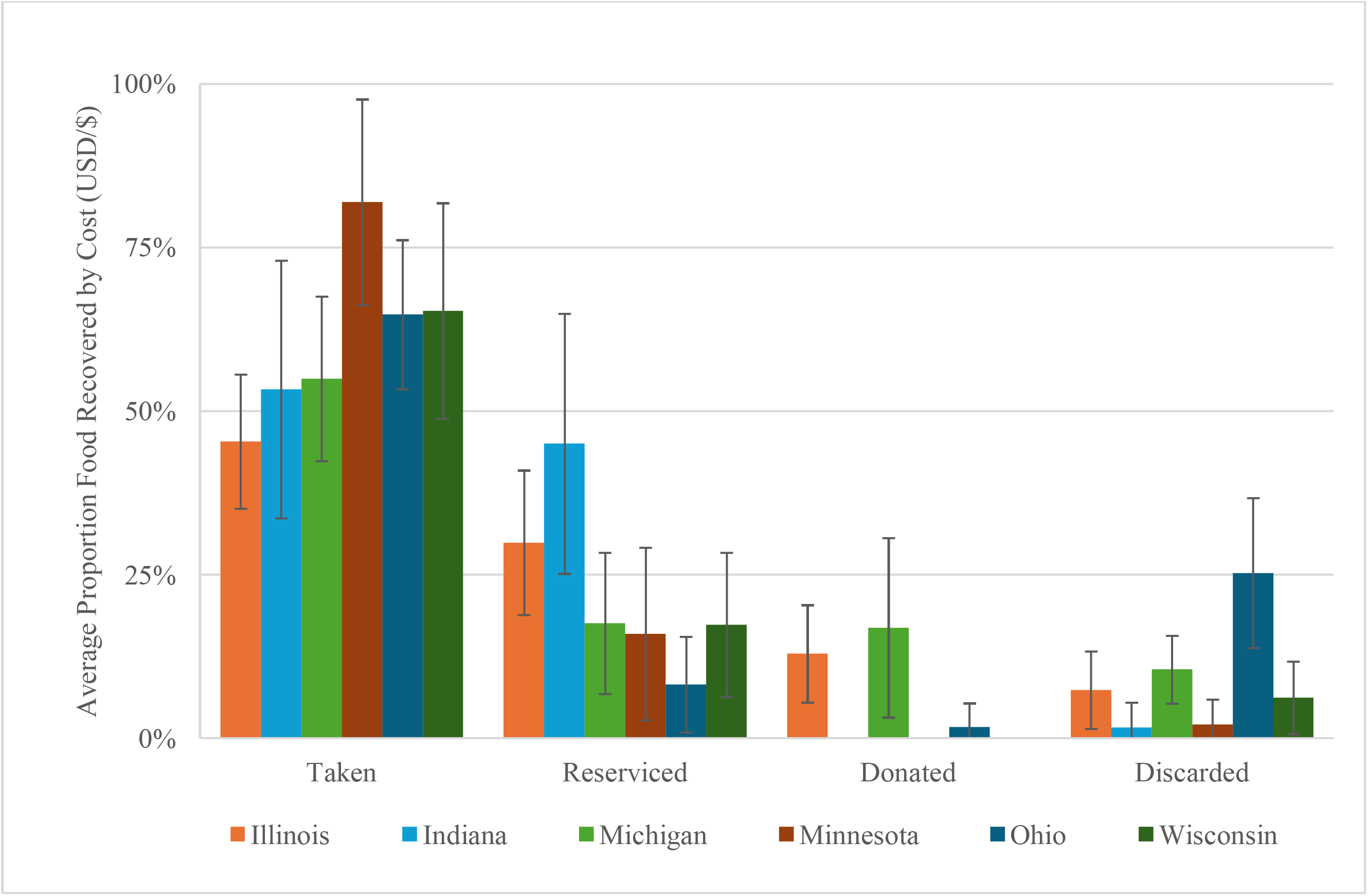
Cost (USD) of foods placed on share tables during observation meals by final use and state

Table 3 provides projections for individual schools across each state to predict the potential amount of food recovered (weight and costs) for the entire school year if a school were to implement a share table at their school. We estimated that, on average, each school in these 6 midwestern states would recover 978 kg (95% PI: 957, 1000) and recover $2,598 (95% PI: $2,565, $2,632) per school in the first year of share table operation with $624 (95% PI: 608, 640) in direct food recovery costs from foods being reserviced. Notably, however, projected food recovery and cost savings were variable across participating states ranging from 324 kg (or $688) per school in Minnesota to 1566 kg (or $4,566) per school in Illinois. Discussion This study estimated the amount and cost of food recovered after implementing school share tables in 41 schools across 6 states, reaching approximately 11,938 students each school day. We concluded that school share tables recover an estimated 978 kg and $2,598 per school in the first year of share table operation. These estimates are solely based on the food students placed on the share table and excludes all uneaten food items that students threw in the trash instead of using the share table. Subsequently, our estimates likely represent the benefit floor of share table food recovery, as student use of the share table is likely to increase over time as students and staff become more accustomed to it. The value of the recovered food does not represent the true fiscal cost of the recovered food because some food purchases are subsidized by the federal government and thus, cost less than market value. Another key finding is that only one school of 41 reported an increase in the number of staff hours worked due to share table implementation, and there was an average of $43 across four schools in the entire pilot study for reported operational expenses. Taken together, these findings suggest that share tables are an inexpensive intervention that improves access to healthy food, reduces food waste, and reduces school nutrition program food costs.

**Table 3.**
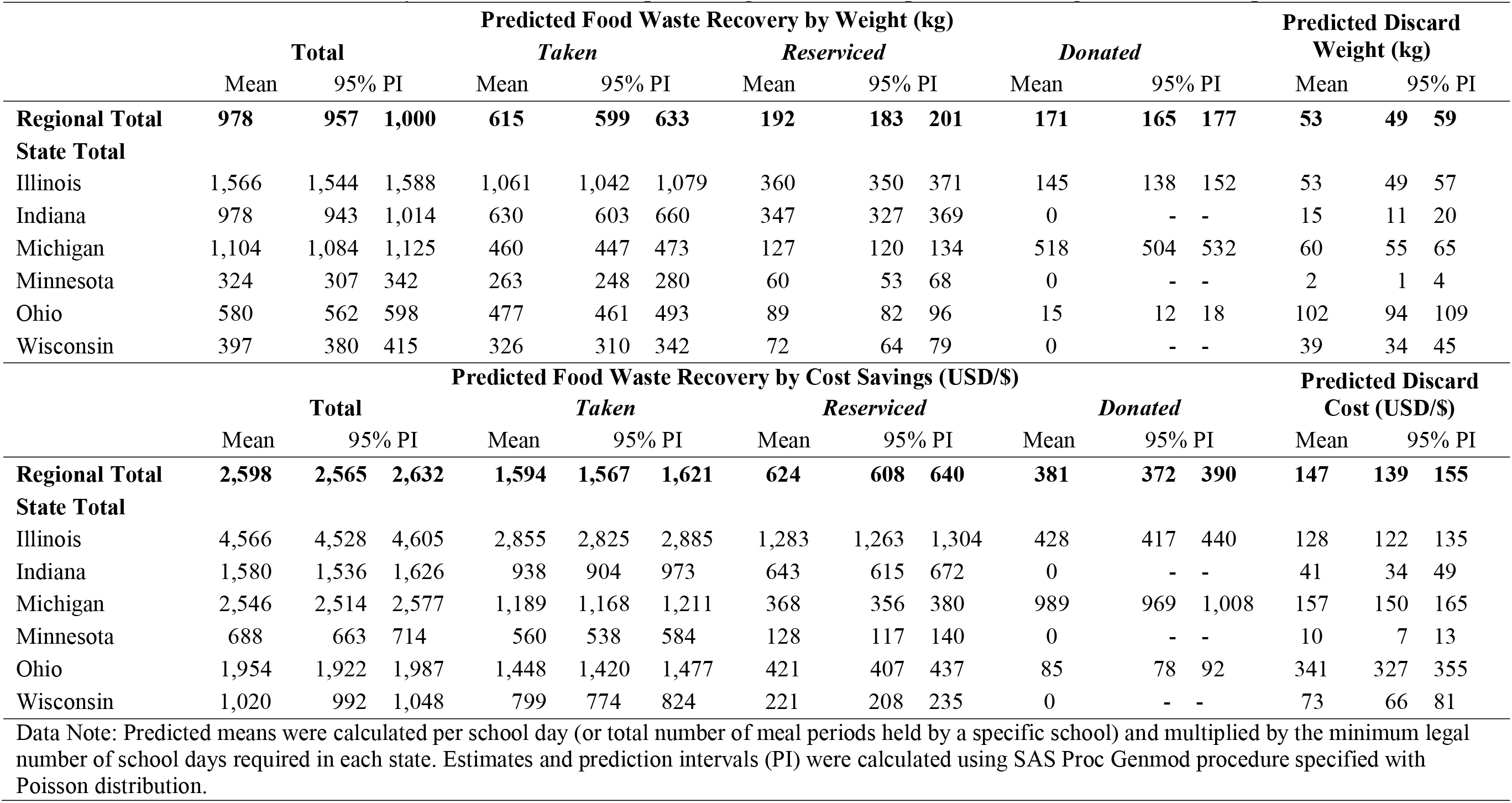
Predicted Mean Food Recovery Metrics Per School Implementing a Share Table per School during First Year of Implementation.

|  | Predicted Food Waste Recovery by Weight (kg) |  |  |  |  |  |  |  |  |  |  |  | Predicted Discard Weight (kg) |  |  |
| --- | --- | --- | --- | --- | --- | --- | --- | --- | --- | --- | --- | --- | --- | --- | --- |
|  | Total |  |  | Taken |  |  | Reserviced |  |  | Donated |  |  |  |  |  |
|  | Mean | 95% PI |  | Mean | 95% PI |  | Mean | 95% PI |  | Mean | 95% PI |  | Mean | 95% PI |  |
| **Regional Total** | **978** | **957** | **1,000** | **615** | **599** | **633** | **192** | **183** | **201** | **171** | **165** | **177** | **53** | **49** | **59** |
| **State Total** |  |  |  |  |  |  |  |  |  |  |  |  |  |  |  |
| Illinois | 1,566 | 1,544 | 1,588 | 1,061 | 1,042 | 1,079 | 360 | 350 | 371 | 145 | 138 | 152 | 53 | 49 | 57 |
| Indiana | 978 | 943 | 1,014 | 630 | 603 | 660 | 347 | 327 | 369 | 0 | - | - | 15 | 11 | 20 |
| Michigan | 1,104 | 1,084 | 1,125 | 460 | 447 | 473 | 127 | 120 | 134 | 518 | 504 | 532 | 60 | 55 | 65 |
| Minnesota | 324 | 307 | 342 | 263 | 248 | 280 | 60 | 53 | 68 | 0 | - | - | 2 | 1 | 4 |
| Ohio | 580 | 562 | 598 | 477 | 461 | 493 | 89 | 82 | 96 | 15 | 12 | 18 | 102 | 94 | 109 |
| Wisconsin | 397 | 380 | 415 | 326 | 310 | 342 | 72 | 64 | 79 | 0 | - | - | 39 | 34 | 45 |
| | Predicted Food Waste Recovery by Cost Savings (USD/\$) | | | | | | | | | | | | Predicted Discard Cost (USD/\$) | | |
|  | Total |  |  | Taken |  |  | Reserviced |  |  | Donated |  |  |  |  |  |
|  | Mean | 95% PI |  | Mean | 95% PI |  | Mean | 95% PI |  | Mean | 95% PI |  | Mean | 95% PI |  |
| **Regional Total** | **2,598** | **2,565** | **2,632** | **1,594** | **1,567** | **1,621** | **624** | **608** | **640** | **381** | **372** | **390** | **147** | **139** | **155** |
| **State Total** |  |  |  |  |  |  |  |  |  |  |  |  |  |  |  |
| Illinois | 4,566 | 4,528 | 4,605 | 2,855 | 2,825 | 2,885 | 1,283 | 1,263 | 1,304 | 428 | 417 | 440 | 128 | 122 | 135 |
| Indiana | 1,580 | 1,536 | 1,626 | 938 | 904 | 973 | 643 | 615 | 672 | 0 | - | - | 41 | 34 | 49 |
| Michigan | 2,546 | 2,514 | 2,577 | 1,189 | 1,168 | 1,211 | 368 | 356 | 380 | 989 | 969 | 1,008 | 157 | 150 | 165 |
| Minnesota | 688 | 663 | 714 | 560 | 538 | 584 | 128 | 117 | 140 | 0 | - | - | 10 | 7 | 13 |
| Ohio | 1,954 | 1,922 | 1,987 | 1,448 | 1,420 | 1,477 | 421 | 407 | 437 | 85 | 78 | 92 | 341 | 327 | 355 |
| Wisconsin | 1,020 | 992 | 1,048 | 799 | 774 | 824 | 221 | 208 | 235 | 0 | - | - | 73 | 66 | 81 |

Data Note: Predicted means were calculated per school day (or total number of meal periods held by a specific school) and multiplied by the minimum legal number of school days required in each state. Estimates and prediction intervals (PI) were calculated using SAS Proc Genmod procedure specified with Poisson distribution.

Research suggests that significant reductions in food waste are one of the top 3 actions necessary to mitigate climate change and otherwise keep the food system within environmental limits. Given climate change’s negative impact on public health, public health agencies and advocates have multiple health priorities to consider related to regulating and administering school share tables: climate change, access to healthy foods, and food safety. Our study findings suggest that share tables have the potential to play a significant role in food waste mitigation and provide an opportunity to divert healthy food from landfills to hungry people. Yet, the types and amount of foods that can be recovered depends upon the perceived food safety of share table operations. Fruit with edible peels and milk are some of the most commonly recovered share table items, but not all health departments allow these foods to be recovered. Norovirus, the most likely food borne illness to stem from share tables, is not fatal. Yet, food sharing is a common practice in school cafeterias outside of share table operation, and school outbreaks of norovirus have occurred prior to share tables being in existence.

While share tables to do present modest food safety risks, research suggests that these risks can be mitigated by improved hand hygiene, washing fruit with edible peels before re-use, Share Only tables, and preserving the cold chain of TCS foods, particularly during overnight storage.^11,14^ In fact, implementing one of the mitigation strategies, hand sanitizing stations prior to entering the meal service line, not only mitigates the norovirus risk of share table operation but also reduces the risk of norovirus transmission of normal cafeteria operations.^11^ Simple mitigation steps, like a cooler with ice or a tray with ice or icepacks, reduces milk spoilage on share tables. The feasibility of share table food safety risk mitigation supports the use of risk-based, rather than hazards-based, approaches to food safety. Hazard-based approaches result in prohibiting food recovery practices because the presence of a potentially harmful virus or other agent exists.^15^ For example, health departments may not allow fruits with edible peels (e.g. apples) to be reserviced when they are leftover on school share tables at the end of lunch service because there could potentially be norovirus particles on the apples after being handled by students. In contrast, risk-based approaches establish guidance according to estimates of human exposure to potentially harmful agent and determines whether risk management action is needed.^15^ Using a risk-based approach, a health department would permit leftover apples to be reserved as long as they were washed first to mitigate norovirus risk. Experts conclude that the field should move away from hazard-based and towards risk-based approaches to ensure food safety because a presence of a hazard does not always translate into risks at certain exposure levels.^16^

Share tables that have effective food safety risk management provide opportunities for safe, healthy food to be diverted from landfills to feed hungry people. This is consistent with the EPA’s Food Recovery Hierarchy, which guides the sustainable reuse of food. In addition, share table foods that are made available during the lunch period or through school backpack programs provide youth with personal agency to obtain free, healthy foods for themselves, and, in some cases, others in their household. This is important because research shows that household participation in federal nutrition assistance programs (e.g., SNAP) does not always translate into improvements in adolescent food security^17^ and the nutrition density of dietary patterns of SNAP-participating households is subpar. This underscores the potential for share tables to promote improved nutrition security among under resourced individuals. The USDA permits unclaimed share table items to be donated externally to food pantries and other non-profits. These donations would fall under the choose often (green) category of the Healthy Eating Research Nutrition Guidelines for the Charitable Food System,^18^ providing a second mechanism for school food recovery to positively impact nutrition security. Furthermore, share tables provide an experiential learning opportunity for students that has the potential to normalize safe food recovery, with potential spillover effects that may benefit future health outcomes through improved food utilization (a component of nutrition security) and climate outcomes over a lifetime of food waste mitigation.

This is the first study to systematically assess the impact of share tables on school food recovery and estimate maintenance costs, but there are important limitations to consider. Most schools who reported additional maintenance costs were those who received the check participation incentive instead of the share table kit. This suggests that these schools may have included purchases they made with their check to establish their share table as maintenance costs. It is also possible that their purchases did not fully anticipate their needs, while the kit was designed to comprehensively meet implementation needs. In addition, we were unable to explore differences between urban and rural schools as only Minnesota and Ohio had nearly equal representation of rural-urban counties. All other participating states had schools that were predominantly rural or predominantly urban. We cannot assume that all schools follow similar patterns of adoption as the schools participating in this pilot; thus, our projections may not be generalizable to all U.S. schools. Our findings are likely most generalizable to other schools with high percentages of free and reduced price lunch eligibility and less generalizable to more affluent schools. The latter are likely to have less food items taken during the lunch period and more opportunities for reservice, donation or landfill. Most of the schools in our sample implemented Share and Take tables, and we were unable to assess differences between share table type. Lastly, there were only 41 schools in our sample which limits our generalizability and power to do more robust analyses.

### Public Health Implications

These study findings show that share tables are an effective strategy to prevent healthy food from being landfilled and can play a key role in climate change mitigation and contribute to a sustainable food system. Maintenance costs of school share tables were low, and the average value of recovered food exceeds the costs of any materials needed to establish a share table, suggesting the potential for widespread adoption and maintenance of share tables across the United States. Share tables also provide important opportunities to improve nutrition security.

These environmental and social benefits of share tables underscore the need for a risk-based approach to food safety. More research and outreach are needed to identify best practices for promoting risk-based approaches to share table regulation to health departments to fully realize the potential of share tables to positively impact public health through reduced greenhouse gas emissions and improved access to healthy foods.

## Data Availability

All data produced in the present study are available for non-commercial purposes upon reasonable request to the authors.

